# Percutaneous Coronary Intervention of Chronic Total Occlusion is Associated with Higher Inpatient Mortality and Complications Compared to Non-CTO lesions Using the National Inpatient Sample (NIS) Database

**DOI:** 10.1101/2023.04.26.23289182

**Authors:** Allistair Nathan, Mehrtash Hashemzadeh, Mohammad Reza Movahed

**Author notes:** Correspondent: M Reza Movahed, MD, PhD, FACP, FACC, FSCAI, FCCP, Clinical Professor of Medicine, University of Arizona Sarver Heart Center, Tucson, Clinical Professor of Medicine, University of Arizona, Phoenix, 1501 N Campbell Avenue Tucson, AZ 85724. **Funding:** None.

## Abstract

**Background:** Percutaneous coronary intervention (PCI) in patients with chronic total occlusion (CTO) is commonly performed despite unclear long-term benefits. The goal of this study was to evaluate the post-procedural outcome of patients with CTO intervention.

**Methods:** The National Inpatient Sample (NIS) database, years 2016-2020, was studied using ICD 10 codes. Patients with CTO intervention were compared to patients without CTO. We evaluated post-procedural mortality and complications.

**Results:** PCI in patients with CTO was associated with higher mortality and all post-procedural complications. A total of 2,011,854 patients underwent PCI with 259,574 having CTO. The CTO group had a 3.17% mortality rate vs 2.57% of non-CTO PCI. (OR, 1.24; CI:1.18–1.31; p<0.001). Using multivariate analysis adjusting for age, sex, race, diabetes mellitus, and chronic kidney disease, CTO PCI remained significantly associated with higher mortality (OR, 1.37; 95% CI, 1.3 – 1.45; p<0.001). Patients with CTO compared to non-CTO PCI patients had higher rates of myocardial infarction (OR, 2.85; 95% CI, 2.54 – 3.21; p<0.001), coronary perforation (OR, 6.01; 95% CI, 5.25-6.89; p<0.001), tamponade (OR, 3.36; 95% CI, 2.91-3.88, p<0.001), contrast-induced nephropathy (OR, 2.05; 95% CI, 1.45-2.90), p<0.001), procedural bleeding (OR, 3.57; 95% CI, 3.27-3.89, p<0.001), and acute post-procedural respiratory failure (OR, 2.07; 95% CI, 1.81-2.36, p<0.001). All post-procedural complications were more than 3 times the non-CTO patients (OR, 3.45; 95% CI, 3.24-3.67; p<0.001).

**Conclusion:** Using a large national inpatient database, PCI performed in patients with CTO was associated with significantly much higher mortality and post-procedural complications compared to PCI in non-CTO patients.

## Introduction

Chronic total occlusion of a coronary artery is defined as a complete or near complete (TIMI 0-1) blockage of flow that has been present for greater than 3 months.^1,2^ Chronic total occlusion has previously been reported to be present in as many as 52% of patients with significant CAD who undergo percutaneous coronary intervention (PCI).^3^ As catheterization technology and revascularization approaches have improved, the number of patients with CTO who undergo PCI has also increased.^4–6^ One 2013 study found that 30% of patients with CTO underwent PCI.^7^

Benefits of PCI in patients with CTO have been reported to include angina relief and improvement in quality of life.^8,9^ Additionally, successful CTO-PCI has been reported to reduce risk of sudden cardiac death, the risk of stroke, and the need for subsequent CABG.^10,11^ However, many of these studies compare successful PCI for CTO with non-successful PCI for CTO, rather than comparing PCI for CTO with optimal medical therapy (OMT). When outcomes from PCI for CTO are compared with those of no-PCI or OMT for CTO, the benefit of PCI in such patients does not exist with no improvement in mortality or hard outcome..^12,13^

Additionally, CTO PCI has been shown to be associated with many potential complications, which may often go underreported.^14^ Reported complications include side branch occlusion, thrombus, coronary artery perforation, MI, arrhythmias, death, and many others.^15^ In particular, the rate of coronary artery perforation has previously been reported to be much higher in CTO-PCI when compared to normal PCI. For example, one 2021 study found coronary artery perforation to occur in <1% of normal PCI cases, however in 4-9% of CTO-PCIs.^16^ Beyond complications, CTO-PCI has also previously been reported to fail at a high rate – one study reported a 24.9% first-attempt failure rate in patients undergoing CTO-PCI.^17^ In line with the conflicting bodies of evidence regarding the procedure’s outcomes, the current guidelines for the use of PCI in the treatment of CTO are vague.^18^

In order to evaluate the true prevalence of in-hospital mortality and adverse events of patients undergoing CTO versus non-CTO PCI, we used a large National Inpatient Sample (NIS) database.

## Methods

### Data Source

The study cohort was derived from the National Inpatient Sample (NIS), Healthcare Cost and Utilization Project (HCUP), and Agency for Healthcare Research and Quality. The NIS database includes discharge information for 2,011,854 patients (10,059,269 after discharge weights), approximating a 20% stratified sample of discharges from U.S. community hospitals, and is representative of 98% of the total U.S. population.^19^ NIS HCUP data is publicly available and deidentified, and thus the study was exempt from institutional review board approval.

### Study Population

Patient data was drawn from the 2016-2020 database years. Both International Classification of Diseases, Tenth Revision, Clinical Modification (ICD-10-CM) and International Classification of Diseases, Tenth Revision, Procedure Coding System (ICD-10-PCS) codes were used to query the NIS database and develop the study cohort. The target population of patients having undergone PCI was identified using the ICD-10-PCS codes 02703(4-7)Z, 02703(D-G)Z, 02703TZ, 02713(4-7)Z, 02713(D-G)Z, 02713TZ, 02723(4-7)Z, 02723(D-G)Z, 02723TZ, 02733(4-7)Z, 02733(D-G)Z, 02733TZ, 02H(0-3)3DZ, 02H(0-3)3YZ, 027(0-3)3ZZ, 02C(0-3)3Z7, 02C(0-3)3ZZ, 02F(0-3)3ZZ. This population was further stratified using the ICD-10-CM code I25.82 to identify patients with CTO. We evaluated patients undergoing PCI with age limits of over 30 years old. Cohort demographic data was calculated using age, gender, and race.

### Study Outcomes

The patient outcomes examined included mortality and complications such as myocardial infarction (MI) (I97.89), contrast-induced nephropathy (N99.0), cardiac perforation (I97.51), procedural bleeding (I97.410, I97.411, I97.610, I97.611, I97.630, I97.631), cardiac tamponade (I31.4), acute postprocedural respiratory failure (J95.821), postprocedural cerebrovascular infarction (I97.821), and major adverse cardiac events (MACE). The MACE outcome was derived from all measured cardiac outcomes as well as mortality. In multivariate analysis, we adjusted mortality for comorbid conditions such as diabetes (ICD-10-CM codes: E10, E11, E13), chronic kidney disease (CKD) (ICD-10-CM codes: I1311, I132, N289, Q613, N181, N182, N183, N1830, N1831, N1832, N184, N185, N186, N189, N19), age, gender, and race.

### Statistical Analysis

Patient demographic, clinical, and hospital characteristics are reported as means, with 95% confidence intervals for continuous variables and proportions, and 95% confidence intervals for categorical variables. Trend analysis over time was assessed using Chi-squared analysis for categorical outcomes and univariate linear regression for continuous variables. Multivariable logistic regression ascertained the odds of binary clinical outcomes relative to patient and hospital characteristics as well as the odds of clinical outcomes over time. All analyses were conducted following the implementation of population discharge weights. All p-values are 2-sided and p<0.05 was considered statistically significant. Data were analyzed using STATA 17 (Stata Corporation, College Station, TX).

## Results

A total of 2,011,854 adult patients were identified who underwent PCI from 2016-2020, and 259,575 of these patients had CTO. The average patient age was 70.04 years (CI: 69.98 – 70.04), and more men (63.68%) underwent PCI than women (36.32%). Caucasian patients composed the majority (77.03%) of the study cohort (*Table 1*).

**Table 1:** Study Cohort Demographics

| **Demographic** | **Cohort Makeup (%)** |
| --- | --- |
| Male | 63.68 |
| Female | 36.32 |
| White | 77.03 |
| Black | 10.6 |
| Hispanic | 7.09 |
| Asian/Pacific Islander | 2.13 |
| Native American | 0.52 |
| Other | 2.63 |
| **Total Patients Studied** | 2,011,854 |
| **Patients with CTO** | 259,575 |

Univariate analysis showed that, in patients who underwent PCI, mortality was significantly higher in patients with CTO than in those without (odds ratio [OR], 1.24; 95% confidence interval [CI], 1.18–1.31; p<0.001). Post-procedural complications were also significantly higher in patients with CTO. Patients with CTO had higher rates of MI (OR, 2.85; 95% CI, 2.54 – 3.21; p<0.001), cardiac perforation (OR, 6.01; 95% CI, 5.25-6.89; p<0.001), cardiac tamponade (OR, 3.36; 95% CI, 2.91-3.88, p<0.001), and MACE (OR, 1.73; 95% CI, 1.66-1.80; p<0.001).

Additionally, CTO PCI was associated with higher rates of contrast-induced nephropathy (OR, 2.05; 95% CI, 1.45-2.90), p<0.001), procedural bleeding (OR, 3.57; 95% CI, 3.27-3.89, p<0.001), and acute post-procedural respiratory failure (OR, 2.07; 95% CI, 1.81-2.36, p<0.001). Total post-procedural complications were significantly higher in CTO PCI (OR, 3.45; 95% CI, 3.24-3.67; p<0.001). These findings are summarized in *Table 2* and *Figure 1*.

**Table 2:** Prevalence of Mortality and Morbidity in Patients with CTO vs. No CTO

| Outcome | Prevalence (%) |  | Odds Ratio (CI) | p-value |
| --- | --- | --- | --- | --- |
|  | CTO | No CTO |  |  |
| Mortality | 3.17 | 2.57 | 1.24 (1.18 - 1.31) | <0.001 |
| Myocardial Infarction | 0.65 | 0.23 | 2.85 (2.54 - 3.21) | <0.001 |
| Contrast-Induced Nephropathy | 0.08 | 0.04 | 2.05 (1.45 - 2.90) | <0.001 |
| Cardiac Perforation | 0.62 | 0.10 | 6.01 (5.25 - 6.89) | <0.001 |
| Procedural Bleeding | 1.12 | 0.32 | 3.57 (3.27 - 3.89) | <0.001 |
| Cardiac Tamponade | 0.4 | 0.12 | 3.36 (2.91 - 3.88) | <0.001 |
| Acute Postprocedural Resp. Failure | 0.46 | 0.22 | 2.07 (1.81 - 2.36) | <0.001 |
| Postprocedural Cerebrovascular Infarction | 0.03 | 0.02 | 1.71 (1.02 - 2.84) | 0.04 |
| MACE | 5.51 | 3.27 | 1.73 (1.66 - 1.80) | <0.001 |
| All Postprocedural Complications | 2.56 | 0.76 | 3.45 (3.24 - 3.67) | <0.001 |

**Figure 1.**
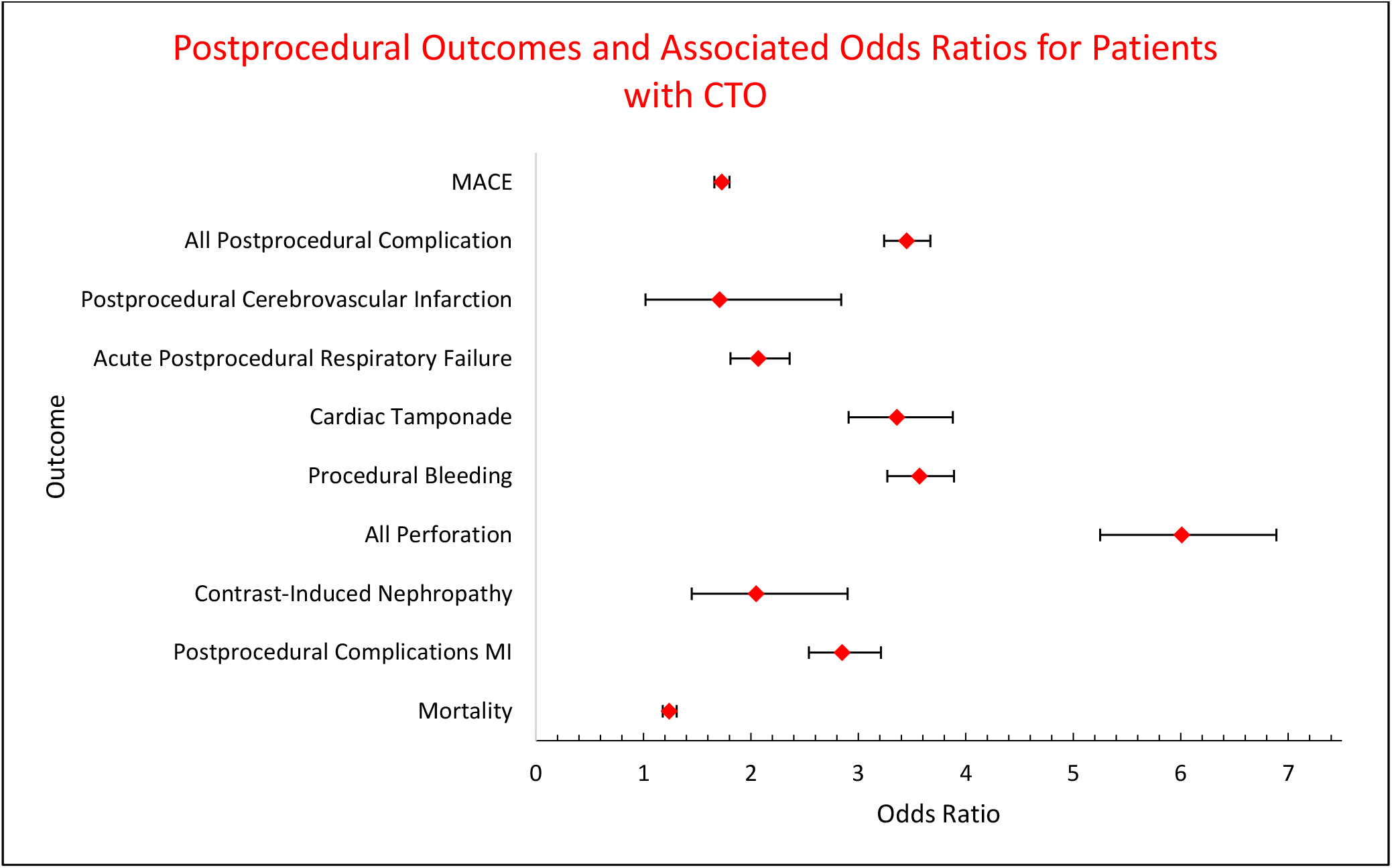
Post-procedural complications were more likely in patients with CTO than without CTO following PCI.

Additionally, CTO remained associated with a significant increase in the risk of mortality following a multivariate analysis adjusting for age, sex, race, diabetes mellitus, and chronic kidney disease (OR, 1.37; 95% CI, 1.3 – 1.45; p<0.001).

## Discussion

Currently, clinical guidelines regarding the use of PCI in patients with CTO are vague. The 2021 American College of Cardiology/American Heart Association Guideline for Coronary Artery Revascularization classified its recommendation of PCI in patients with CTO as “weak”.^18^ Such inconclusive guidelines have been fueled by conflicting bodies of evidence regarding the utility of PCI for CTO. For example, in their 2005 retrospective analysis, Stone et al. conclude that given PCI success rates of 80-90% in patients with CTO, PCI should be the preferred method of treatment.^20^ On the other hand, the EXPLORE randomized-controlled trial found no benefit associated with PCI when compared to optimal medical treatment in the management of patients with CTO, as measured by left ventricular function.^21^ Similar results were found in the REVASC trial, which used segmental wall thickening in the CTO territory as its primary end-point.^22^ Additionally, the DECISION-CTO trial, which enrolled over 800 patients, demonstrated no significant difference in outcomes between PCI and optimal medical treatment in CTO patients.^12^

Considering these randomized controlled trials demonstrate no significant benefit associated with CTO-PCI, the uncertainty regarding the efficacy of the procedure may be fueled by studies comparing successful vs unsuccessful CTO-PCI. These studies, such as that conducted by Xenogiannis et al., often conclude that PCI for CTO leads to favorable outcomes with lower complication rates.^23^ However, comparing successful CTO-PCI to non-successful CTO-PCI has been reported to be inherently flawed.^24^ For example, Xenogiannis et al. report successful CTO-PCI leading to lower rates of angina and MACE.^23^ However, it is important to note that this conclusion of decreased morbidity is dependent upon the increase in morbidity observed in the unsuccessful PCI study arm. Any of the adverse events observed in the unsuccessful arm would not have occurred had the patients been given OMT, as was the case in the previously discussed randomized trials. Additionally, such studies often neglect to emphasize the role of baseline patient characteristics (often poorer in the failed CTO PCI group), which was the case in the study conducted by Xenogiannis et al.^25^ Thus, comparing successful and failed CTO-PCI is a flawed means of assessing the efficacy and safety of PCI for CTO, and results from such studies should not be considered when developing guidelines for the use of PCI for CTO.

When the results of our study are considered alongside those of the EXPLORE, REVASC, and DECISION-CTO, the benefit of performing PCI on patients with CTO is brought into question.^12,21,22^ Our study, which is the largest retrospective study comparing the outcomes of PCI in CTO vs no-CTO patients, demonstrates that performing PCI for CTO contributes to significantly higher morbidity and complications. We found that mortality and all measures of morbidity (*Table 2, Figure 1*) were dramatically more prevalent in patients with CTO, emphasizing the much higher risk of PCI in these patients Even upon adjusting for age, sex, race, diabetes, and chronic kidney disease, this study found that mortality was still 37% more likely in the CTO population.

Considering the results of the previously discussed randomized trials and those of the current study, it is clear that PCI for CTO is an unnecessarily risky procedure. In light of this risk, PCI for CTO is performed at an alarmingly high rate in the United States. For example, as stated earlier, our study identified 259,575 patients with CTO who underwent PCI between the years 2016 and 2020, representing 12.9% of all patients who underwent PCI. While CTO-PCI may be appropriate in cases of severe, refractory angina, its current use is rarely limited to such severe cases.^25^ For example, in the previously discussed study by Xenogiannis et al., less than 50% of the patients were on nitrate or calcium channel blockers.^23^ Additionally, a previous study using NIS data from the years 2008-2014 identified 109,094 patients with CTO who underwent PCI.^26^ In their study, Doshi et al. explored trends in CTO-PCI hospitalization rates, in-hospital mortality, acute renal failure, the use of mechanical circulatory support devices, and cost, between the years 2008 and 2014.^26^ The study was not comparative and did not seek to assess the efficacy or safety of PCI for CTO. Not only did their study demonstrate a high rate of CTO-PCI incidence, but it found that PCI utilization in the management of CTO rose 9% from 2008 to 2014.^26^ Doshi et al. also found that the odds of developing post-procedural acute renal failure increased between 2008 and 2014, as did the use of percutaneous LVADs and the cost of hospitalization.^26^ Thus, this study conducted by Doshi et al. not only emphasizes the overuse of CTO-PCI but compliments our findings in suggesting the use of PCI in patients with CTO contributes to an avoidable financial burden placed on our healthcare system, which should be studied further.

Considering our study’s findings and those of the previous studies mentioned, a significant body of evidence now supports the use of optimal medical therapy in patients with CTO rather than PCI, which clinical guidelines should begin to reflect. CTO-PCI should only be performed in severely symptomatic patients with symptoms resistant to maximal medical therapy. Unfortunately, CTO-PCI is too commonly performed with poor indication, endangering many patients.

## Data Availability

NIS data available to the public

